# Pre-conception PfSPZ Vaccine administration to Malian women prevents malaria in their offspring during infancy

**DOI:** 10.64898/2026.11.19.26368501

**Authors:** Halimatou Diawara, Sara A. Healy, Jing Wang, Zonghui Hu, Djibrilla Issiaka, Seydou Traore, Ibrahim H. Soumbounou, Santara Gaoussou, Almahamoudou Mahamar, Oumar Attaher, Ibrahim Diarra, Natasha KC, Tooba Murshedkar, Viyada Doan, Aye Diallo, Robert D. Morrison, Justin Y.A. Doritchamou, B. Kim Lee Sim, Thomas L. Richie, Stephen L. Hoffman, Alassane Dicko, Patrick E. Duffy

**Affiliations:** Parasites and Microbes Training and Research Center, University of Sciences, Techniques and Technologies of Bamako, Mali; Laboratory of Malaria Immunology and Vaccinology, National Institute of Allergy and Infectious Diseases, National Institutes of Health, Bethesda, MD, USA; Clinical Monitoring Research Program Directorate, Frederick National Laboratory for Cancer Research, Frederick, MD, USA; Biostatistical Research Branch, National Institute of Allergy and Infectious Diseases, National Institutes of Health, Bethesda, MD, USA; Sanaria, Inc., Rockville, MD, USA

## Abstract

**Background:** Preconception administration of *Plasmodium falciparum* sporozoite (PfSPZ) Vaccine is safe and prevents malaria during pregnancies identified within 21 months post-vaccination. We evaluated pre-conception PfSPZ Vaccine effects on malaria risk in offspring during the first year of life.

**Methods:** In a randomized, double-blind, placebo-controlled trial in Ouélessébougou, Mali, 300 healthy 18-38-year-old women were allocated 1:1:1 to receive 9×10^5^ PfSPZ or 1.8×10^6^ PfSPZ of PfSPZ Vaccine or normal saline on days 1-8-29, and followed up to 21 months for malaria endpoints and pregnancies. 104 infants from 103 pregnancies identified first year post-vaccination [9×10^5^ PfSPZ (n=38); 1.8×10^6^ PfSPZ (n=40); normal saline (n=26)] were seen every two months and during illnesses. Primary endpoints were blood-smear parasitemia and clinical malaria. Safety outcomes included newborn baseline characteristics and infant adverse events (AEs).

**Findings:** Newborn baseline characteristics and infant AEs did not differ between arms; AEs were common but predominantly mild (91.9–96% Grade 1). Vaccine efficacy of 9×10⁵ PfSPZ was 75.1% (95% CI 20.6–92.2) against parasitemia (10.5% parasitemic infants versus 38.5% of placebo) and 94.2% (54.9–99.3) against clinical malaria (2.6% malarious infants versus 38.5% of placebo). VE of 1.8×10^6^ PfSPZ group was 40.3% (−43.6-75.2) against parasitemia and 36.3% (−50.3-73.0) against clinical malaria, both non-significant. At 6 and 12 months of age, PfCSP antibodies were similar in offspring of vaccinated and control mothers.

**Interpretation:** Pre-conception PfSPZ Vaccine was safe and reduced Pf infection and clinical malaria during infancy in 9×10⁵ PfSPZ group. Further development of preconception PfSPZ vaccines is warranted to protect mother-infant dyads.

## INTRODUCTION

Malaria is the leading cause of morbidity and mortality in sub-Saharan Africa. More malaria cases and deaths occurred in 2024 than 2014,^1^ and under-5 children carry the heaviest burdens despite wide deployment of the use of insecticide treated nets (ITN) and seasonal malaria chemoprevention (SMC). While infants under 6 months enjoy reduced malaria risk, presumably due to maternal antibodies, risk progressively increases ^2,3^ and malaria is common during later infancy.^4–6^ New tools that protect infants include R21 or RTS,S vaccines administered as 3 monthly doses starting age 5 months or later.

*Plasmodium falciparum* (Pf) sporozoite (SPZ) vaccines are in advanced development for use in pregnant women, owing in part to highly favorable safety profiles.^7^ In a study of radiation-attenuated Sanaria^®^ PfSPZ Vaccine launched in Ouélessébougou, Mali in 2019, 300 non-pregnant women anticipating pregnancy in the next year were allocated 1:1:1 to receive 9.0×10^5^ PfSPZ, 1.8×10^6^ PfSPZ or normal saline. PfSPZ Vaccine showed no safety signals and reduced parasitemia and clinical malaria, including during pregnancies identified during 21 months of post-vaccination follow up.^8^

In this study, newborns of women who became pregnant within 1 year of last PfSPZ Vaccine dose were followed through infancy. Adverse event, parasitemia, and clinical malaria data were collected every 2 months at routine clinical visits, as well as during any illness. We examined impact of maternal vaccine assignment on offspring safety and efficacy outcomes.

## METHODS

### Ethical considerations

The trial followed International Council for Harmonization Good Clinical Practice guidelines and Malian regulations. Community permission from the village chief and elders and individual written informed consent was obtained from all participants. Trial protocols were approved by ethics committees of the US National Institutes of Health (Bethesda, MD, USA), the Faculté de Médecine et d’Odonto-stomatologie (Bamako, Mali), and Mali Ministry of Health (Bamako, Mali). Studies were conducted with the US Food and Drug Administration (FDA) Investigational New Drug Application. The trial was registered at clinicaltrials.gov (NCT03989102).

### Study design and participants

The study was conducted at the Ouélessébougou Clinical Research Unit, Parasites and Microbes Research and Training Center (PMRTC), located 80 km south of Bamako, Mali. Malaria transmission is highly seasonal occurring between July and December, with 1.99 clinical malaria episodes per under-five child per year.^9^

In the main study, conducted 2019 to 2023, women received artemether-lumefantrine to clear malaria parasitemia, then were randomly allocated 1:1:1 to receive PfSPZ Vaccine at 9×10^5^ or 1.8×10^6^ PfSPZ per dose, or normal saline placebo, at weeks 0, 1 and 4, using double-blind design. Women received contraception from one month before first vaccination until 1 month after 3^rd^ vaccination, then discontinued at their discretion. Pregnancy was identified by monthly hCG testing and followed through delivery. All pregnant women were encouraged to receive intermittent preventive treatment in pregnancy (IPTp) starting from second trimester, per Mali Ministry of Health standard of care. More details are in the main trial publication.^8^

For pregnancies confirmed by hCG within 1 year after the 3^rd^ and last vaccine or placebo administration, women were invited to enroll their newborn for 12 months post-delivery follow up. Analyses excluded infants from mothers who did not receive 3 vaccine or placebo administrations. Newborns underwent first examination at birth including anthropometric measurements, with CBC and blood-smear microscopy of placenta and cord blood samples. Newborns were actively assessed every 2 months during infancy, and passively through availability of the study clinicians 24 hours a day, seven days a week to assess any medical complaints during infancy. Blood-smear and adverse event data were collected at each scheduled and unscheduled visit; blood was collected at 6 and 12 months of age for immunology studies. Women were encouraged to independently bring children for routine EPI and seasonal malaria chemoprevention (SMC) delivered through Ministry of Health.

### Outcomes

Baseline newborn characteristics were assessed by maternal vaccine arm, including infant gender and birth season [low (January–June) or high malaria transmission (July– December)]. For safety outcomes, we compared proportions of adverse events, including birth outcomes, between infants from 9×10^5^ PfSPZ, 1.8×10^6^ PfSPZ, and placebo groups. Efficacy outcomes were infant parasitemia and clinical malaria, and we estimated vaccine efficacy by maternal vaccine versus comparator arm for both parasitemia and clinical malaria.

*P. falciparum* (Pf) blood stage infection was defined as at least 2 asexual Pf parasites detected in 0.5 µL blood on Giemsa-stained smear. Consistent with Mali National Malaria Control Programme guidelines, clinical (symptomatic) malaria was defined as presence of any asexualLmalariaLparasites and/or positive rapid diagnostic test with either axillary temperature 37.5 °C or more, and/or one or more of the following: history of fever, chills, headache, myalgia, arthralgia, nausea, vomiting, diarrhea, and abdominal pain. Clinical malaria was reported as an AE. Low birth weight was defined as <2500 grams per WHO guidelines.^10^

IgG antibodies to P. falciparum circumsporozoite protein (PfCSP) were measured in sera from mothers at delivery and from infants at 6 and 12 months of age, by ELISA as described previously.^8,11,12^ Antibodies to merozoite surface protein-1 (PfMSP1) were assessed in the same sera using previously described ELISA methods.^13–15^ T-cell subsets (including Vδ2^+^) were enumerated on freshly collected blood using previously described methods.^8^

### Statistical analysis

We performed per protocol (PP) analyses on 104 infants. Differences in proportions of AEs between vaccine groups and controls were assessed by Fisher’s exact test. Vaccine efficacies against parasitemia and clinical malaria were primarily evaluated using time-to-event analyses, with efficacy estimated as one minus the hazard ratio from a Weibull proportional hazards model for interval-censored data (using function icweib in R package “straweib”). This model compared each vaccine arm with placebo and, in separate sensitivity analysis, incorporated delivery season (low/high transmission) and gravidity as stratification factors or covariates. Additional analyses assessed proportions experiencing at least one event, with efficacy estimated as one minus the relative risk using exact binomial test. Differences between groups in antibody assays were assessed using Mann-Whitney U test.

Post-hoc sensitivity analyses, using updated statistical tools and measures while accounting for additional variables to improve rigor, are detailed in **Supplementary Appendix**.

## RESULTS

The 300 main trial participants received last dose of vaccine from 07-Aug-2019 to 27-Aug-2019. In total, 118 infants were born to 114 women whose pregnancy was identified within a year of last vaccine dose, including 3 sets of twins. First year pregnancies, documented by hCG testing, occurred from 23-Oct-2019 to 15-Sep-2020 for 9×10^5^ PfSPZ vaccine arm; 29-Oct-2019 to 15-Sep-2020 for 1.8×10^6^ PfSPZ; 20-Aug-2019 to 22-Sep-2020 for placebo. Deliveries occurred 17-Jun-2020 to 10-May-2021 for 9×10^5^ PfSPZ vaccine arm; 22-Jun-2020 to 09-May-2021 for 1.8×10^6^ PfSPZ; 30-Apr-2020 to 17-May-2021 for placebo.

Among these 118 infants, 104 were included in this analysis. 14 infants were excluded due to either incomplete maternal vaccination (N=2), withdrawn consent (N=8), or neonatal death (N=4). Neonatal deaths included two sets of twins delivered preterm at 27- and 28-weeks gestational age. The 104 infants (103 pregnancies), including 1 set of twins, were followed from birth to age 12 months between 30-Apr-2020 until 5-Apr-2022: from 18-Jun-2020 to 16-Mar-2022 for 9×10^5^ PfSPZ vaccine arm (N=38); 22-Jun-2020 to 16-Mar-2022 for 1.8×10^6^ PfSPZ (N=40); 30-Apr-2020 to 05-Apr-2022 for placebo (N=26).

The proportion of female offspring was roughly half and did not differ significantly based on maternal vaccine assignment (**Table 1**). More children were born during malaria transmission season, but proportions did not differ significantly between treatment arms, ranging from 57.7-68.4%. Mean birth weight and length were similar across treatment arms.

**Table 1.**
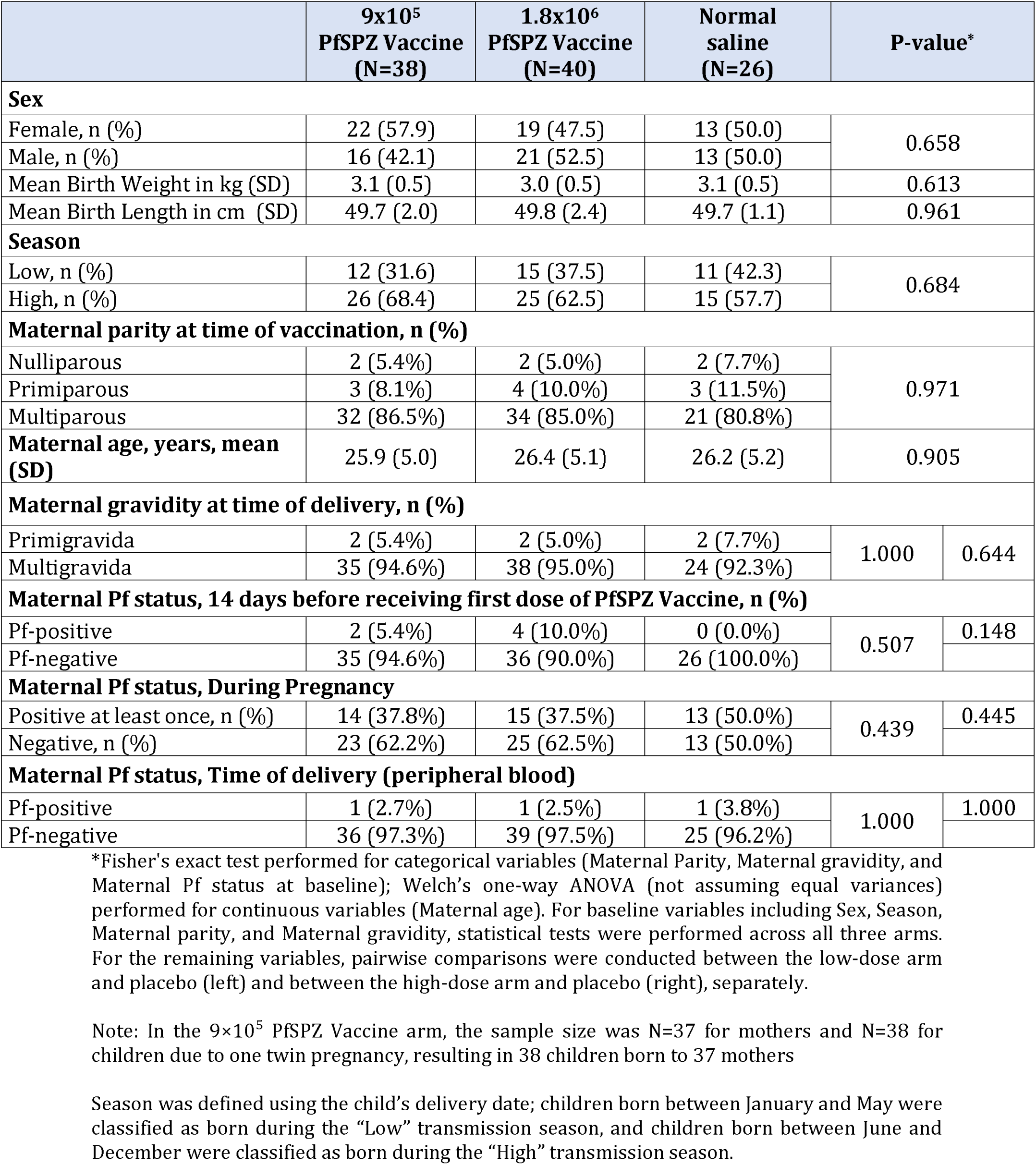
Baseline information of infants by mother vaccine arm.

### Safety

During follow up, proportion of total AEs was similar between infants of all arms, and the majority were mild in severity (**Supplementary Table S1**). Commonest AEs included rhinitis and respiratory infections for all arms, with the large majority mild (grade 1), including 95.8% in 9×10^5^ PfSPZ Vaccine arm, 92.7% in 1.8×10^6^ arm, and 93.2% in placebo arm (**Table 2**). Seventeen serious adverse events (SAEs) were reported in 15 infants (**Supplementary Tables S2**), including 5 (13.2%) in the 9×10^5^ PfSPZ arm (3 premature birth, 2 bronchiolitis, and 1 neonatal infection), 8 (20.0%) in 1.8×10^6^ PfSPZ arm (3 premature birth, 2 fetal distress, 1 neonatal infection, 1 respiratory infection, and 1 malaria), and 2 (7.7%) in the placebo arm (1 fetal distress, 1 bronchiolitis, and 1 malnutrition). No significant differences were observed between treatment groups for any or all SAEs, nor for poor pregnancy outcomes (low birth weight, preterm delivery, and small for gestational age; **Table 3**), as well as cord and placental CBC by either vaccine group or infection outcome (**Supplementary Tables S3-S4**).

**Table 2.** Adverse events during first year of life by mother vaccine arm.

| **All AEs** | **9x10<sup>5</sup><br>PfSPZ Vaccine<br>(N=38)** | **1.8x10<sup>6</sup><br>PfSPZ Vaccine<br>(N=40)** | **Normal saline<br>(N=26)** | **P-value<br>(Low dose vs.<br>Normal saline)** | **P-value<br>(High dose vs.<br>Normal saline)** |
|---|---|---|---|---|---|
| Rhinitis | 18 (47.4%) 25 | 22 (55.0%) 35 | 14 (53.8%) 20 | 0.799 | 1.000 |
| Respiratory tract infection | 18 (47.4%) 28 | 19 (47.5%) 24 | 14 (53.8%) 20 | 0.799 | 0.801 |
| Sinobronchitis | 15 (39.5%) 18 | 18 (45.0%) 24 | 12 (46.2%) 15 | 0.617 | 1.000 |
| Gastroenteritis | 14 (36.8%) 23 | 11 (27.5%) 13 | 10 (38.5%) 12 | 1.000 | 0.422 |
| Clinical Malaria | 1 (2.6%) 1 | 11 (27.5%) 12 | 10 (38.5%) 17 | 0.0003 | 0.422 |
| **Others** | 33 (86.8%) 97 | 34 (87.5%) 98 | 20 (76.9%) 64 | 0.331 | 0.517 |
| **Total AEs** | 36 (94.7%) 192 | 39 (97.5%) 206 | 26 (100%) 148 |  |  |

Numbers reported are unique individuals experiencing a specific AE (percentage of individuals who experienced the specific AE) total number of that specific AE.
The AE of clinical (symptomatic) malaria was defined as a positive blood smear or rapid diagnostic test with either axillary te mperature 37.5 °C or more, and/or one or more of the following: history of fever, chills, headache, myalgia, arthralgia, nausea, vomiting, diarrhea, and a bdominal pain.
Differences between groups were analyzed by Fisher's exact test based on the number of individuals with a specific AE.

**Table 3.** Birth Outcomes.

|  | **9x10<sup>5</sup><br>PfSPZ Vaccine<br>(N=38)** | **1.8x10<sup>6</sup><br>PfSPZ Vaccine<br>(N=40)** | **Normal<br>Saline<br>(N = 26)** | **P-value<br>(Low dose vs.<br>Normal saline)** | **P-value<br>(High dose vs.<br>Normal saline)** |
|---|---|---|---|---|---|
| **Low Birth Weight (<2.5 kg)** | 3 (7.9%) | 3 (7.5%) | 0 (0.0%) | 0.265 | 0.273 |
| **Preterm (<37 weeks GA)** | 3 (7.9%) | 4 (10.0%) | 3 (11.5%) | 0.680 | 1.000 |
| **Small for Gestational Age** | 2 (5.3%) | 3 (7.5%) | 0 (0.0%) | 0.510 | 0.273 |

Numbers in table are number of infants as percentage of total infants within each treatment group
<sup>2</sup>Fisher's Exact Test

### Vaccine efficacy

#### Protocol-defined analyses

Primary VE analysis assessed time to first Pf infection (i.e., positive blood-smear), and each vaccine arm was compared separately to placebo. The 9×10^5^ PfSPZ Vaccine arm conferred significant protective efficacy against Pf infection for infants [VE=75.1% (20.6-92.2), p=0.016; **Figure 1A**, **Table 4**]. The 1.8×10^6^ PfSPZ Vaccine arm conferred nonsignificant VE of 40.3% (95%CI: −43.6-75.2, p=0.248). No non-falciparum episodes occurred.

**Figure 1.**
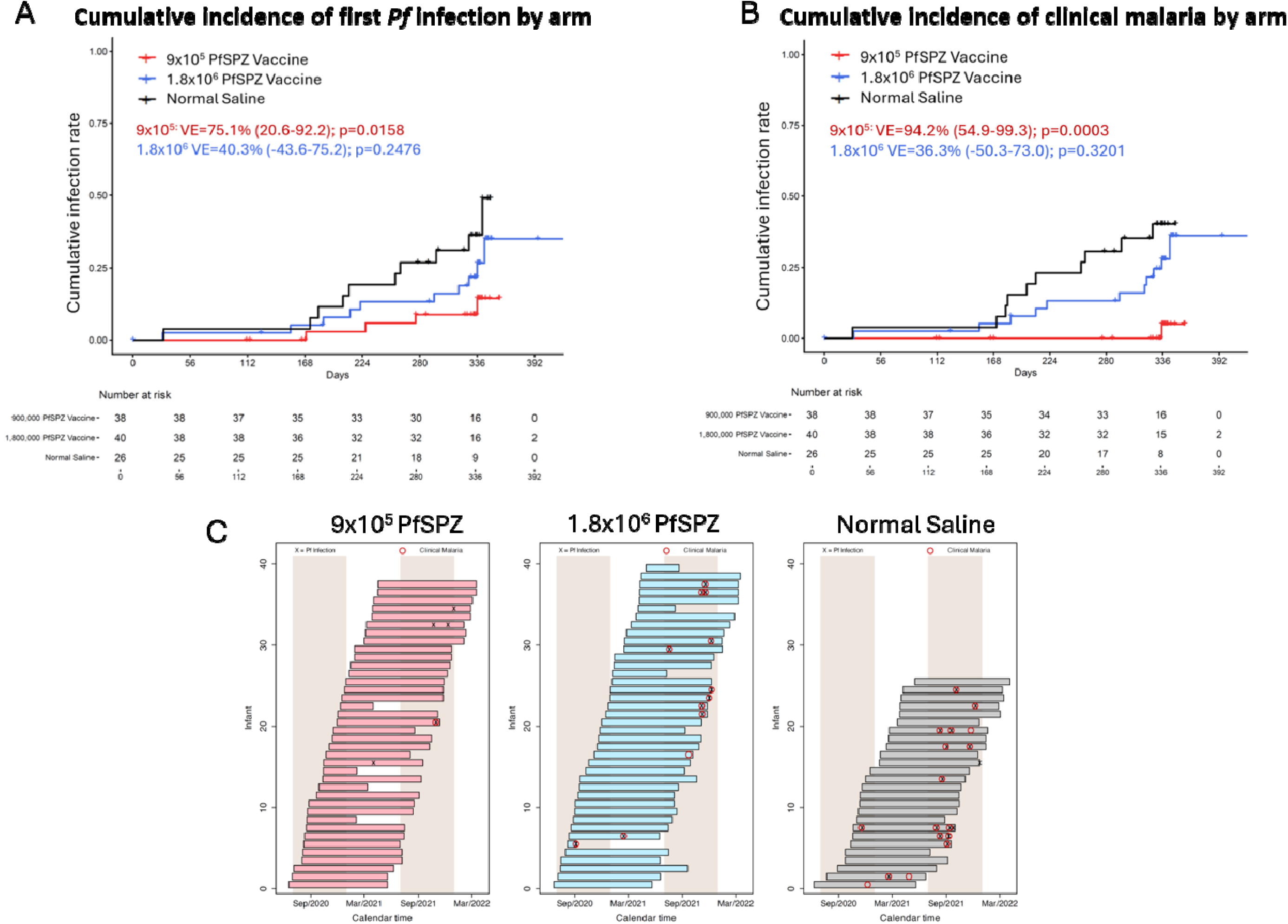
Cumulative incidence of first *Plasmodium falciparum* (Pf) infection and clinical malaria by arm. Kaplan–Meier curves show the cumulative incidence of A) first Pf infection, and B) first clinical malaria episode over the follow-up period for offspring of women receiving 9×10^5^ PfSPZ Vaccine, 1.8×10^6^ PfSPZ Vaccine, or normal saline. Tick marks indicate censored observations, and the numbers at risk are displayed below the x-axis. Vaccine efficacy (VE) and corresponding 95% confidence intervals (CI) are indicated on the plot, as well as P values from interval-censored log-rank tests comparing each vaccine group with the normal saline group. Positive VE values indicate a lower hazard of first Pf infection or first clinical malaria compared with the normal saline group. C) Swimmer plots showing malaria events during infancy by study arm assignment to 9×10^5^ PfSPZ Vaccine, 1.8×10^6^ PfSPZ Vaccine, or normal saline detailing per protocol study population (defined as offspring of women who received all three vaccinations as randomized). Each row represents offspring of women who became pregnant within a year of last vaccine dose. X symbol indicates Pf parasitemia detected by blood-smear, and red circle indicates clinical malaria episode; a red circle that does not contain an X indicates clinical malaria diagnosed by RDT without a positive blood-smear; two parasitemia episodes within 7 days are indicated as a single X symbol at the time of first positive blood-smear. Tan background shading indicates malaria transmission seasons (July to December).

**Table 4:** Vaccine efficacy against first *Plasmodium falciparum* infection in infants during first year of life. Vaccine efficacy (VE) was estimated as one minus the hazard ratio from a proportional hazards model for interval-censored data assuming a Weibull baseline survival function. Each vaccine group was compared separately with the normal saline control group. 95% confidence intervals (CIs) were calculated from the fitted model, and P values were obtained from interval-censored log-rank tests. Positive VE values indicate a lower hazard of first *P. falciparum* infection compared with the normal saline control group.

| **Comparison** | **VE<br>(%)** | **95% CI<br>(Lower)** | **95% CI<br>(Upper)** | **P value<br>(Log-rank Test)** |
|---|---|---|---|---|
| 9x10<sup>5</sup> PfSPZ Vaccine vs. Normal Saline | 75.12 | 20.61 | 92.20 | 0.016 |
| 1.8x10<sup>6</sup> PfSPZ Vaccine vs. Normal Saline | 40.30 | -43.59 | 75.18 | 0.248 |

The secondary VE analysis assessed time to first clinical malaria episode. The 9×10^5^ PfSPZ Vaccine arm again conferred consistent and significant efficacy against clinical malaria (VE=94.2% (54.9-99.3), p=0.0003; **Figure 1B**, **Table 5**). Through day 336, none of 38 infants in 9×10⁵ PfSPZ group had clinical malaria, compared with 10 of 26 placebo infants (**Figure 1B**). The 1.8×10^6^ PfSPZ Vaccine arm conferred nonsignificant VE of 36.3% (95%CI :-50.3-73.0, p=0.320).

**Table 5:** Vaccine efficacy against first clinical malaria in infants during first year of life. Vaccine efficacy (VE) was estimated as one minus the hazard ratio from a proportional hazards model for interval-censored data assuming a Weibull baseline survival function. Each vaccine group was compared separately with the normal saline control group. 95% confidence intervals (CIs) were calculated from the fitted model, and P values were obtained from interval-censored log-rank tests. Positive VE values indicate a lower hazard of first clinical malaria compared with the normal saline control group.

| **Comparison** | **VE<br>(%)** | **95% CI<br>(Lower)** | **95% CI<br>(Upper)** | **P value<br>(Log-rank Test)** |
|---|---|---|---|---|
| 9x10<sup>5</sup> PfSPZ Vaccine vs. Normal Saline | 94.23 | 54.90 | 99.26 | 0.0003 |
| 1.8x10<sup>6</sup> PfSPZ Vaccine vs. Normal Saline | 36.26 | -50.25 | 72.96 | 0.320 |

Parasitemia (27/32) and clinical malaria (24/30) events occurred primarily during second malaria season, and second half of infancy (26/32 and 26/30), respectively (**Figure 1C**). In placebo arm, 1/16 parasitemia episodes occurred in first malaria season when children were almost exclusively under-6-month-olds (7-12-month-old mean exposure, 1 day), 1/16 in dry season (mean exposure, 43 days), and 14/16 in second malaria season (mean exposure, 90 days).

Both primary and secondary VE results were supported by additional protocol-defined analyses based on proportion of participants, a stratified interval-censored proportional hazard model, and an interval-censored proportional hazard model adjusted for birth season. Each analysis demonstrated significant efficacy against first Pf infection (**Tables S5-S7**) and clinical malaria (**Tables S8-S10**) conferred by 9×10^5^ PfSPZ dose, but not 1.8×10^6^ PfSPZ dose.

Additional sensitivity analyses (details in **Appendix; Tables S11-S18**) confirmed protocol-defined analyses: that 9×10^5^ PfSPZ Vaccine conferred consistent and significant protection against Pf parasitemia and clinical malaria in infants of vaccinated mothers, while 1.8×10^6^ PfSPZ Vaccine dose did not. We found no significant association between protection in the mother during pregnancy and protection in the infant (**Supplemental Results; Table S19**), albeit with limited sample size.

### Immunology Assays

PfCSP is the major surface protein of PfSPZ and target of protective antibodies induced by RTS,S and R21 or given by passive transfer.^16^ At delivery, vaccinated mothers had significantly higher anti-PfCSP IgG than mothers given normal saline (median OD 1.0: 908 for 9×10^5^ PfSPZ; 1,189 for 1.8×10^6^ PfSPZ and 344 for normal saline; p<0.0001, Mann-Whitney U test) (**Figure 2; Table S20**), albeit a >6-fold decrease from peak post-vaccination titers (5,773) for 9×10^5^ PfSPZ mothers.^17^ Infant anti-PfCSP IgG at 6 and 12 months was low (highest OD 1.0 of 80; in 9 × 10⁵ PfSPZ group) and did not differ between arms. In 9×10⁵ PfSPZ group, mother delivery anti-PfCSP IgG was higher when the infant remained uninfected throughout follow up (median OD 1.0: 943, n=30 vs. 336, n=4 infected; p=0.042). No other comparison by infection status was significant. Secondary analyses that split month 0–6 and 7–12 infections gave similar results, including the significant 9×10⁵ PfSPZ group findings (**Supplementary Figure S1**).

**Figure 2.**
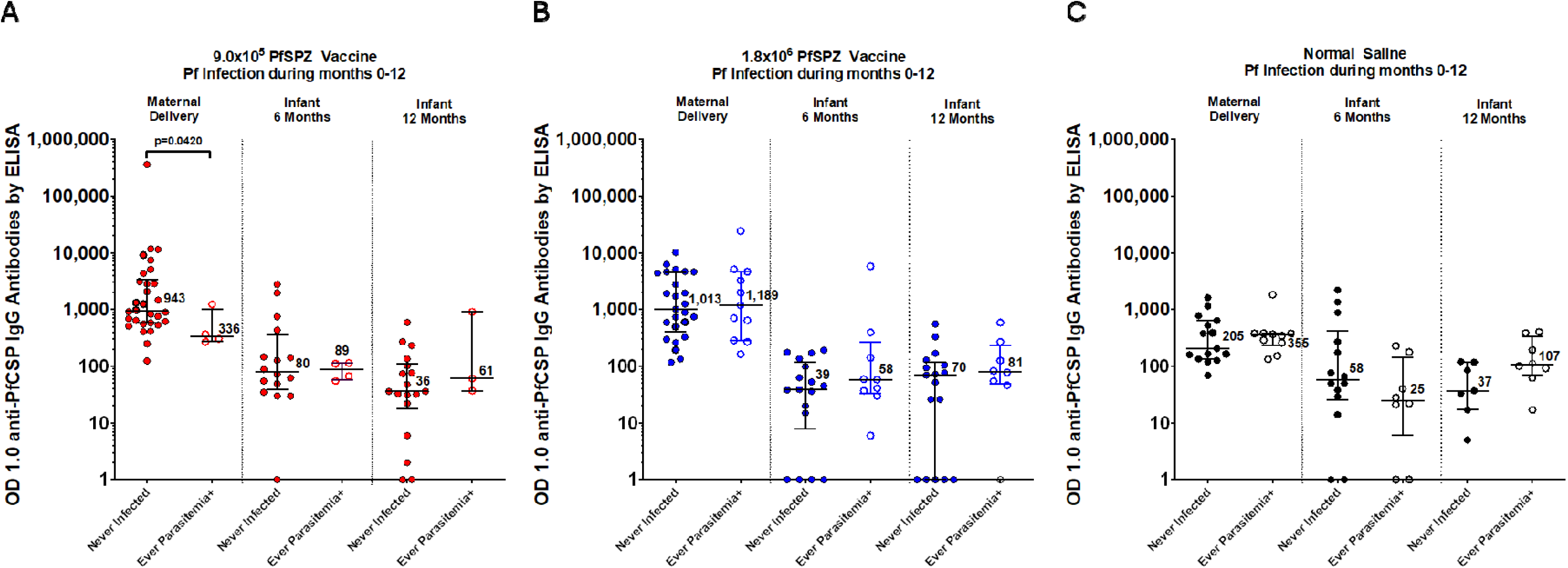
PfCSP-specific IgG antibody responses measured by ELISA in vaccinated mothers at delivery and in their infants at 6 and 12 months of age. Data are presented as the median and interquartile range of the reciprocal serum dilution at which the optical density was 1.0 (OD 1.0) and are shown separately for participants receiving 9.0×10⁵ PfSPZ Vaccine, 1.8×10⁶ PfSPZ Vaccine, and normal saline. Within each panel, responses are stratified by infant infection status during the corresponding follow-up period. Each symbol represents an individual participant; horizontal lines indicate medians, and error bars represent interquartile ranges. OD 1.0 values are displayed on a log₁₀ scale. Of the 104 infants followed (38 in 9×10^5^; 40 in 1.8×10^6^; 26 in the control group) 34/38, 37/40 and 25/26 of the mothers had sera available at the time of delivery, 20/38, 26/40, and 22/26 of the infants had sera available at 6 months and 21/38, 25/40, and 15/26 of the infants had sera available at 12 months. Differences between groups were assessed using the Mann-Whitney U test. The p-values were not corrected for multiple comparisons.

PfMSP1 is a blood-stage antigen. Maternally transferred anti-PfMSP1 antibodies have associated with protection from clinical malaria during infancy,^15,16^ while malaria-resistant children show PfMSP1 antibody seroprevalence.^17^ At delivery, anti-PfMSP1 IgG antibodies did not differ between mothers given 9×10^5^ PfSPZ, 1.8×10^6^ PfSPZ, and normal saline (median EUs: 628, 481, and 453). Median infant levels at 6 and 12 months were near limit of detection in all arms (**Figure 3; Table S20**); previously parasitemic infants had much higher EUs than uninfected infants in every arm (median 261 vs. 3 EU, 9×10⁵ PfSPZ (p=0.007); 1,403 vs. 3 EU, 1.8×10⁶ (p=0.0005); 927 vs. 3 EU, normal saline (p=0.001)). The secondary analysis by age-period was similar (Supplementary Figure S2).

**Figure 3.**
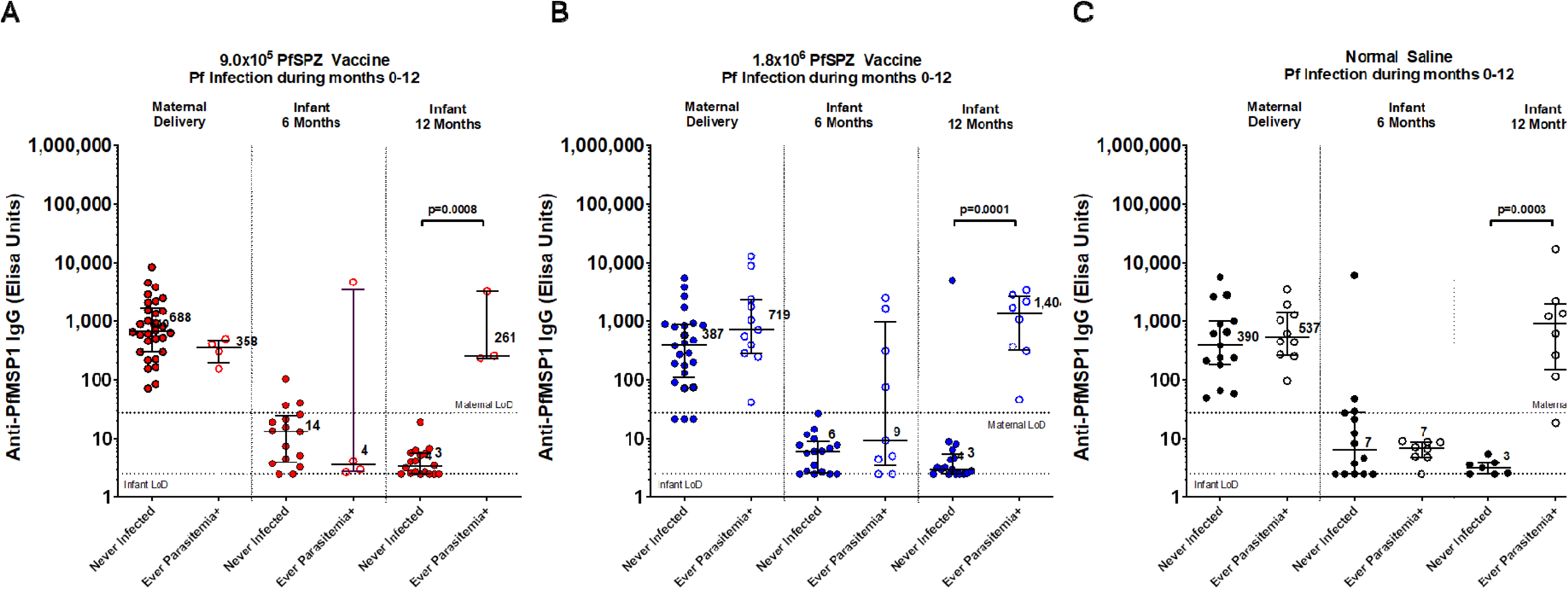
PfMSP1-specific IgG antibody responses measured by ELISA in vaccinated mothers at delivery and in their infants at 6 and 12 months of age. Data are presented as the median and interquartile range of anti-PfMSP1 ELISA units (EU) and are shown separately for participants receiving 9.0 × 10⁵ PfSPZ Vaccine, 1.8 × 10⁶ PfSPZ Vaccine, and normal saline. Within each panel, responses are stratified by infant infection status during the corresponding follow-up period. Each symbol represents an individual participant; horizontal lines indicate medians, and error bars represent interquartile ranges. ELISA units are displayed on a log₁₀ scale. The horizontal reference lines indicate the respective PfMSP1 ELISA limit-of-detection (LoD) seropositive thresholds for maternal and infant samples. Of the 104 infants followed (38 in 9×10^5^; 40 in 1.8×10^6^; 26 in the control group) 34/38, 37/40 and 25/26 of the mothers had sera available at the time of delivery, 20/38, 26/40, and 22/26 of the infants had sera available at 6 months and 21/38, 25/40, and 15/26 of the infants had sera available at 12 months. Differences between groups were assessed using the Mann-Whitney U test. The p-values were not corrected for multiple comparisons.

In this and prior PfSPZ Vaccine trials, fold-increase in Vδ2 γδT-cells during and after vaccination has been associated with protection, and γδ T-cells are essential to generate protective liver CD8^+^ T-cells in SPZ-immunized rodents.^18^ We did not observe significant differences between treatment arms, nor between infected and uninfected infants, in 1) Vδ2 levels in maternal delivery or infant samples (**Figure S3**), 2) Vδ2 foldchange in infant samples from maternal delivery sample baseline (**Figure S4**).

## DISCUSSION

In this trial, preconception PfSPZ Vaccine was associated with 94.2% reduction in clinical malaria incidence among infants of vaccinated mothers. This finding extends our previous observation that pregnancy malaria risk decreased 57% in the 9×10^5^ PfSPZ arm.^8^ If confirmed, these results would establish preconception vaccination as an approach to protect both women and their future offspring from malaria, without requiring vaccination of infants.

Infant protection may be related to PfSPZ Vaccine efficacy in the mother. While pregnancy malaria (PM) has been associated with increased infection risk of offspring in several studies, including our trial site,^19–21^ this relationship is confounded by shared exposure. In this trial, randomization removed confounding factors, enabling assessment of whether vaccine-induced protection against PM can also reduce risk in their offspring. In the main trial, 9×10^5^ PfSPZ Vaccine conferred greater efficacy against pregnancy malaria than 1.8 × 10^6^ PfSPZ Vaccine (57% vs 49%),^7^ as also observed in Tanzanian adults (100% vs 33%),^22^ although mechanisms remain unknown. Since SPZ vaccine efficacy in animals is mediated by liver tissue-resident CD8+ T cells,^23–29^ higher antigen doses might tolerize or suppress these responses.

While PfSPZ Vaccine protection has been imputed mainly to CD8+ T cells that recognize Pf-infected hepatocytes,^25^ passively transferred maternal immunity against other pathogenns is typically attributed to transplacental IgG. At delivery, maternal PfCSP IgG levels were highest in 9×10^5^ PfSPZ arm, whose offspring remained largely free of Pf infection throughout infancy. However, titers were minimal and did not differ between arms at 6 or 12 months of age (**Figure 2**), spanning the period when nearly all parasitemia events occurred (**Figure 1C**). The paucity of malaria events in first transmission season is consistent with resistance in under-6-month-olds due to passively transferred maternal IgG, whereas the paucity in dry season suggests second malaria season parasitemias was related to new exposures rather than recrudescences.

Based on our findings, we speculate that durable protection in infants, like their mothers, was mediated by T cells, a concept supported by small animal and nonhuman primate studies of whole-sporozoite vaccination.^25,30–33^ Transfer and persistence of maternal immune cells through maternal microchimerism^34^ or altered fetal immune programming resulting from malaria exposure during pregnancy^35^ could contribute, though neither was investigated in this trial.

Pre-conception vaccination to protect offspring against malaria or any disease during the second half of infancy is unprecedented. For other diseases, vaccination before pregnancy can benefit infants by preventing congenital anomalies (e.g. rubella, varicella), vertical transmission (e.g. hepatitis B), or pregnancy complications. Pre-conception immunization with rubella vaccine avoids live virus exposure during pregnancy and protects offspring for 2-6 months through maternal transfer of anti-rubella IgG. Likewise, immunization of pregnant women with RSV vaccine during the 3^rd^ trimester protects offspring from birth through 6 months by maternal transfer of anti-RSV IgG.

The results must be interpreted cautiously considering several limitations. This was a single site Phase 2 clinical trial with a modest population (n=104 infants), and analysis was restricted to the per-protocol population. The small sample size limits precision of the efficacy estimate, interpretation of subgroup findings, and assessment of clinical outcomes like prematurity and low birth weight. Despite the limited sample size, the lower limit of 95% confidence interval regarding vaccine efficacy was 55% against clinical malaria.

This study provides evidence that protection conferred by pre-conception PfSPZ Vaccine before, during, and after pregnancy^8^ can extend to protection against malaria in offspring. Preconception vaccination could complement existing childhood interventions: ITN, SMC and licensed vaccines RTS,S and R21 protect young children, and combining RTS,S and SMC further reduces malaria disease and death.^36,37^ However, despite SMC and ITN, 10 of 26 (38.5%) placebo arm infants developed clinical malaria, highlighting the urgent need for new countermeasures. An ongoing trial (NCT07675785) in Mali of next-generation PfSPZ-LARC2 Vaccine^38^ seeks to confirm benefits of preconception vaccination for mother-infant dyads.

## Data Availability

All data produced in the present work are contained in the manuscript and are available upon reasonable request to the authors

## Acknowledgements

This work was supported in part by the Intramural Research Program of the National Institutes of Health, The contributions of the NIH author(s) are considered Works of the United States Government. The findings and conclusions presented in this paper are those of the author(s) and do not necessarily reflect the views of the NIH or the U.S. Department of Health and Human Services. The work was also funded by the National Cancer Institute, National Institutes of Health, under Contract No.75N91019D00024. The authors thank J. Patrick Gorres for assistance in manuscript preparation, Manufacture of PfSPZ Vaccine by Sanaria was funded partly by the NIAID Small Business Innovation Research grant (5R44AI055229–11). Pharmaceutical operations, regulatory and clinical support by Sanaria was supported by Sanaria. The original trial was funded partly by the Grand Challenges Canada’s Saving Lives at Birth programme.

## Notes

### Competing Interest Statement

B. Kim Lee Sim and Stephen L. Hoffman are employees and stockholders of Sanaria, Inc, the manufacturer of PfSPZ Vaccine.

### Clinical Trial

NCT03989102

### Author Declarations

The trial followed International Council for Harmonization Good Clinical Practice guidelines and Malian regulations. We obtained community permission from the village chief and elders and then individual written informed consent from all participants. Trial protocols were approved by ethics committees of the US National Institutes of Health (Bethesda, MD, USA), the Faculte de Medecine et d'Odonto-stomatologie (Bamako, Mali), and Mali Ministry of Health (Bamako, Mali). Studies were conducted with the US Food and Drug Administration (FDA) Investigational New Drug Application. The trial was registered at clinicaltrials.gov (NCT03989102).

