## Supplementary Material for "Pre-conception PfSPZ Vaccine administration to Malian women prevents malaria in their offspring during infancy"

#### Supplementary Appendix

### 1. SUPPLEMENTARY METHODS

#### Cord blood and placental sample collections

Umbilical cord blood and placental specimens were collected at delivery by trained study personnel in Ouelessebougou, Mali. When immediate neonatal resuscitation or emergency care was not required, the umbilical cord was clamped after pulsation had ceased. Following placental delivery, the placenta was placed in a clean plastic bag on a tray, the fetal membranes were removed, and the maternal surface was cleaned with normal saline until gauze applied to the surface remained free of visible blood. With the fetal surface facing upward, approximately 5 mL of cord blood was aspirated from the umbilical vein using a sterile 10-mL syringe fitted with an 18-gauge blunt-ended needle and preloaded with 1–2 mL of 2% ethylenediaminetetraacetic acid (EDTA) in saline. Blood was transferred to a collection tube. Cord length and placental weight were recorded in the electronic case report form.

The placenta was placed in a clean bag containing 3 mL of 2% EDTA in saline. Tissue was sampled from an area of the fetal surface free of large vessels and reserved for fixed and frozen preparations. Placental blood collection proceeded immediately after tissue sampling to minimize clotting. The remaining placenta was cut into approximately 10 × 10-cm pieces and mechanically disrupted using an autoclaved tissue grinder. The collection beaker was preloaded with 5 mL of 2% EDTA in saline, and an additional 1 mL was added for each placental piece during grinding. The resulting placental blood was filtered through a mesh and transferred into 50-mL conical tubes. Blood accumulating separately in the collection bag was aspirated with a syringe and transferred into separate tubes. Tubes were labeled with the study identifier, collection date, and specimen type, distinguishing blood obtained by grinding from blood collected directly from the bag.

For frozen preparations, placental tissue was covered with polyvinyl alcohol freezing medium, wrapped in aluminum foil, placed in a plastic bag, and frozen at −80°C. After removing the portion of placental tissue intended for freezing, the remaining placenta was cut into five pieces and placed into a grinder for processing. For frozen preparations, placental tissue samples were covered with freezing medium (polyvinyl alcohol mix), wrapped in aluminum foil, placed in a plastic bag, and stored at −80°C until shipment to the National Institutes of Health (NIH) for further processing and analysis.

#### Sensitivity Analysis

In addition to the protocol-defined primary efficacy analysis, we further conducted post-hoc sensitivity analyses using updated statistical tools and measures and accounting for additional variables to improve rigor. In a series of sensitivity tests, VE was calculated against both Pf parasitemia and clinical malaria in the following ways:

1) as one minus the hazard ratio which was estimated from a semiparametric proportional hazard model that allows for interval censoring (using R package “icsp2”, an updated version of “icenReg”);
2) at 300 days because the number of participants remaining at risk by Day 365 was limited. Therefore, an earlier time point was selected to provide a more stable estimate of VE. The proportion of infection in each treatment arm was estimated as one minus the Kaplan–Meier estimate, assuming infections occurred at the time they were detected. VE was calculated as one minus the relative risk of infection comparing each vaccine arm with the normal saline arm
3) using separate baseline hazard functions specified for each gravidity stratum while estimating a common treatment effect across strata. Each vaccine group was compared separately with the normal saline group.
4) using an interval-censored proportional hazard model adjusted for birth season

### 2. SUPPLEMENTARY RESULTS

#### Sensitivity Analyses

Given the positive results from the protocol-defined primary efficacy analysis, we further conducted a series of sensitivity analyses using the most up-to-date statistical tools and measures to give more robust, reliable and stable estimates of VE. Additional sensitivity tests accounted for maternal gravidity and birth season. Results of these sensitivity analyses against Pf infection are described and presented in **Tables S11-S14**, and results of the same tests against clinical malaria are presented in **Tables S15-S18**. Overall, the sensitivity analyses confirmed the findings of the protocol-defined analyses: that $9x10^5$ PfSPZ Vaccine conferred consistent and significant protection against Pf parasitemia and clinical malaria in infants of vaccinated mothers, while the $1.8x10^6$ PfSPZ Vaccine dose did not.

#### Relationship between Maternal and Infant Protection

We investigated possible association between protection in the mother during pregnancy and protection in the infant by stratifying infection status during pregnancy and during first year of infancy (**Table S17**). Examining women that received the $9x10^5$ PfSPZ dose (which conferred VE), 16 of 38 mothers who gave birth had parasitemia during pregnancy. Half (2/4) of infants who had Pf infection during first year of life were born to mothers who had Pf infection during pregnancy. Of infants who remained Pf-negative during first 12 months of life 20 of 34 (59%) were born to mothers who had Pf infection during pregnancy. Proportions were similar for both the $9x10^5$ and placebo groups, and no significant association between maternal and infant infectious statuses was attributed by Fisher's exact test, though small sample sizes may have precluded accurate estimates.

#### Vδ2 γδ T cells

Several PfSPZ trials, including the previous trial of pre-conception women,[8] observed significantly higher Vδ2 γδ T cell responses in individuals that remained uninfected vs. those infected, and mouse studies indicate these cells are required to induce protective liver CD8 T-cells.[16] Here, no significant differences in raw Vδ2 percentages were seen between infants born to vaccinated mothers vs. controls, neither at delivery, nor 6 or 12 months of life.

#### 3. SUPPLEMENTARY SAFETY TABLES

**Table S1. Infant adverse events by severity and by mother vaccine arm**

| Severity | $9x10^5$ PfSPZ Vaccine (N=38) | $1.8x10^6$ PfSPZ Vaccine (N=40) | Normal saline (N=26) | P-value |
|---|---|---|---|---|
| Grade 1 - Mild | 36 (94.7%) 184 | 39 (97.5%) 191 | 26 (100%) 138 | 0.620 |
| Grade 2 - Moderate | 4 (10.5%) 5 | 9 (22.5%) 9 | 7 (26.9%) 8 | 0.202 |
| Grade 3 - Severe | 3 (7.9%) 3 | 5 (12.5%)5 | 2 (7.7%)2 | 0.767 |
| Grade 4 - Life-threatening | 0 (0.0%) 0 | 0 (0.0%)0 | 0 (0.0%) 0 | — |
| Grade 5 - Death | 0 (0.0%) 0 | 1 (2.5%)1 | 0 (0.0%) 0 | 1.000 |
| **Total AEs** | **36 (94.7%) 192** | **39 (97.5%) 206** | **26 (100%) 148** | 0.620 |

P-value calculated on unique participant counts using Fisher's exact test

All participants with a grade 2 AE or higher also had at least one grade 1 AE; therefore the number of participants in the Total AE row is identical to that in the Grade 1 row.

**Table S2. All Serious Adverse Events by mother vaccine arm**

| Serious Adverse Event | $9x10^5$ PfSPZ Vaccine (N=38) | $1.8x10^6$ PfSPZ Vaccine (N=40) | Normal saline (N=26) | P-value |
|---|---|---|---|---|
| Premature baby | 3 (7.9%) 3 | 3 (7.5%) 3 | 0 (0.0%) 0 | 0.439 |
| Fetal distress syndrome | 0 (0.0%) 0 | 2 (5.0%) 2 | 1 (3.8%) 1 | 0.466 |
| Bronchitis / Bronchiolitis | 2 (5.3%) 2 | 0 (0.0%) 0 | 1 (3.8%) 1 | 0.354 |
| Neonatal infection | 1 (2.6%) 1 | 1 (2.5%) 1 | 0 (0.0%) 0 | 1.000 |
| Malnutrition | 0 (0.0%) | 0 (0.0%) 0 | 1 (3.8%) 1 | 0.250 |
| Respiratory tract infection | 0 (0.0%) 0 | 1 (2.5%) 1 | 0 (0.0%) 0 | 1.000 |
| Malaria | 0 (0.0%) 0 | 1 (2.5%) 1 | 0 (0.0%) 0 | 1.000 |
| **Total Participants with ≥1 SAE** | **5 (13.2%) 6** | **8 (20.0%) 8** | **2 (7.7%) 3** | **0.431** |

P-value calculated on unique participant counts using Fisher's exact test

**Table S3. Cord blood CBCs**

| Cell Type | **9x10$^5$ PfSPZ** N=33 | **1.8x10$^6$ PfSPZ** N=35 | **Normal Saline** N=25 | **p-value** (Kruskal-Wallis) |
|---|---|---|---|---|
| WBC (10$^3$/UL) | 8.40 (6.96, 9.84) | 9.45 (7.65, 11.24) | 8.27 (6.56, 9.97) | 0.5118 |
| HGB (G/DL) | 11.39 (10.27, 12.52) | 11.53 (10.37, 12.67) | 11.71 (10.77, 12.65) | 0.9257 |
| PLAT (10$^3$/UL) | 215.47 (169.63, 261.31) | 199.08 (168.13, 230.04) | 211.72 (168.28, 255.16) | 0.6602 |
| ANC (10$^3$/UL) | 3.78 (3.07, 4.49) | 4.45 (3.52, 5.38) | 4.19 (3.05, 5.32) | 0.5209 |
| ALC (10$^3$/UL) | 3.54 (2.57, 4.52) | 3.82 (2.81, 4.84) | 2.86 (2.30, 3.42) | 0.3352 |
| **Cell Type** | **Protected** N=66 | **Infected** N=27 | | **p-value** (T test) |
| WBC | 9.15 (7.96, 10.35) | 7.80 (6.40, 9.19) | | 0.1382 |
| HGB | 11.65 (10.96, 12.34) | 11.22 (9.84, 12.6) | | 0.573 |
| PLAT | 208.76 (181.58, 235.93) | 207.16 (165.59, 248.74) | | 0.9481 |
| ANC | 4.30 (3.65, 4.95) | 3.75 (2.93, 4.57) | | 0.2917 |
| ALC | 3.60 (2.92, 4.28) | 3.132 (2.38, 3.89) | | 0.3526 |

Numbers presented are mean values with 95% confidence intervals.

**Table S4. Placental blood CBCs**

| Cell Type | 900K PfSPZ | 1.8M PfSPZ | Normal Saline | p-value (Kruskal-Wallis) |
|---|---|---|---|---|
| WBC (10^3/UL) | 0.07, (-0.06, 0.20) N=38 | 0.01 (-0.01, 0.03) N=40 | 1.55 (-1.62, 4.71) N=26 | 0.6201 |
| HGB (G/DL) | 8.54 (6.57, 10.51) N=35 | 9.70 (6.86, 12.54) N=36 | 8.15 (5.96, 10.35) N=25 | 0.8153 |
| PLAT (10^3/UL) | 6.00 (5.11, 6.89) N=35 | 6.32 (5.29, 7.35) N=37 | 6.98 (6.03, 7.93) N=25 | 0.4341 |
| ANC (10^3/UL) | 341.75 (243.93, 439.58) N=35 | 310.90 (225.06, 396.74) N=36 | 293.76 (161.60, 425.92) N=25 | 0.5524 |
| ALC (10^3/UL) | 3.1 (2.37, 4.07) N=35 | 3.79 (2.65, 4.93) N=36 | 3.79 (2.68, 4.91) N=25 | 0.5086 |
| **Cell Type** | **Protected** | **Infected** | | **p-value** (T test) |
| WBC | 0.04 (-0.02, 0.10) N=77 | 1.48 (-1.56, 4.53) N=27 | | 0.3401 |
| HGB | 8.98 (7.22, 10.74) N=69 | 8.60 (6.62, 10.58) N=27 | | 0.7742 |
| PLAT | 6.62 (5.94, 7.29) N=70 | 5.75 (4.84, 6.67) N=27 | | 0.129 |
| ANC | 325.59 (253.64, 397.54) N=69 | 297.47 (205.98, 388.97) N=27 | | 0.6253 |
| ALC | 3.42 (2.71, 4.14) N=69 | 3.99 (2.92, 5.05) N=27 | | 0.3732 |

Numbers presented are mean values with 95% confidence intervals and sample size.

#### 3.2 PROTOCOL-DEFINED ANALYSES OF VACCINE EFFICACY

**Table S5. Vaccine efficacy based on the proportion of participants with first *Plasmodium falciparum* infection.** The number and proportion of participants with at least one P. falciparum infection are summarized for each treatment group. Each vaccine group was compared separately with the Normal Saline group. Relative risks (RRs) and odds ratios (ORs) were estimated using exact binomial test. Vaccine efficacy (VE) was calculated as one minus the RR and is presented with two-sided 95% confidence intervals (CIs). P values were obtained from exact binomial tests for the RR.

| | # subjects with infection | # subjects in this arm | VE (95% CI) (1 − RR) × 100 | Odds ratio (95% CI) | P-value |
|---|---|---|---|---|---|
| Normal Saline | 10 | 26 | | | |
| $9x10^5$ PfSPZ Vaccine | 4 | 38 | 73 (16, 93) | 0.19 (0.04, 0.80) | 0.0194 |
| $1.8x10^6$ PfSPZ Vaccine | 10 | 40 | 35 (-51, 72) | 0.53 (0.16, 1.79) | 0.3735 |

**Table S6. Vaccine efficacy against first *Plasmodium falciparum* infection estimated using a stratified interval-censored proportional hazard model.** Separate baseline hazard functions were specified for each Gravidity stratum while estimating a common treatment effect across strata. Each vaccine group was compared separately with the Normal Saline group. P values were obtained from Wald tests of the treatment effect from the model.

| Comparison | VE (%) | 95% CI (Lower) | 95% CI (Upper) | P-value (Wald test) |
|---|---|---|---|---|
| $9x10^5$ PfSPZ Vaccine vs. Normal Saline | 79.05 | 28.36 | 93.87 | 0.0127 |
| $1.8x10^6$ PfSPZ Vaccine vs. Normal Saline | 44.74 | -34.94 | 77.37 | 0.1929 |

**Table S7. Vaccine efficacy against first *Plasmodium falciparum* infection estimated using an interval-censored proportional hazard model adjusted for birth season.** Birth season was included in the model as an adjustment covariate. Each vaccine group was compared separately with the Normal Saline group. P values were obtained from Wald tests of the treatment effect from the model.

| Comparison | VE (%) | 95% CI (Lower) | 95% CI (Upper) | P-value (Wald test) |
|---|---|---|---|---|
| $9x10^5$ PfSPZ Vaccine vs. Normal Saline | 74.80 | 19.50 | 92.11 | 0.0200 |
| $1.8x10^6$ PfSPZ Vaccine vs. Normal Saline | 38.48 | -48.87 | 74.58 | 0.2812 |

**Table S8. Vaccine efficacy based on the proportion of participants with clinical malaria.** The number and proportion of participants with at least one clinical malaria are summarized for each treatment group. Each vaccine group was compared separately with the Normal Saline group. Relative risks (RRs) and odds ratios (ORs) were estimated using exact binomial test. Vaccine efficacy (VE) was calculated as one minus the RR and is presented with two-sided 95% confidence intervals (CIs). P values were obtained from exact binomial tests for the RR.

| | # subjects with infection | # subjects in this arm | VE (95% CI) (1 – RR) × 100 | Odds ratio (95% CI) | P-value |
|---|---|---|---|---|---|
| Normal Saline | 10 | 26 | | | |
| 9x10$^5$ PfSPZ Vaccine | 1 | 38 | 93 (56, 100) | 0.04 (0.00, 0.36) | 0.0006 |
| 1.8x10$^6$ PfSPZ Vaccine | 11 | 40 | 29 (-63, 68) | 0.61 (0.18, 2.01) | 0.5049 |

**Table S9. Vaccine efficacy against first clinical malaria estimated using a stratified interval-censored proportional hazards model.** Separate baseline hazard functions were specified for each Gravidity stratum while estimating a common treatment effect across strata. Each vaccine group was compared separately with the Normal Saline group. P values were obtained from Wald tests of the treatment effect from the model.

| Comparison | VE (%) | 95% CI (Lower) | 95% CI (Upper) | P-value (Wald test) |
|---|---|---|---|---|
| 9x10$^5$ PfSPZ Vaccine vs. Normal Saline | 96.43 | 60.81 | 99.67 | 0.0064 |
| 1.8x10$^6$ PfSPZ Vaccine vs. Normal Saline | 39.89 | -43.89 | 74.89 | 0.2531 |

**Table S10. Vaccine efficacy against first clinical malaria estimated using an interval-censored proportional hazards model adjusted for birth season.** Birth season was included in the model as an adjustment covariate. Each vaccine group was compared separately with the Normal Saline group. P values were obtained from Wald tests of the treatment effect from the model.

| Comparison | VE (%) | 95% CI (Lower) | 95% CI (Upper) | P-value (Wald test) |
|---|---|---|---|---|
| 9x10$^5$ PfSPZ Vaccine vs. Normal Saline | 94.22 | 54.77 | 99.26 | 0.0066 |
| 1.8x10$^6$ PfSPZ Vaccine vs. Normal Saline | 34.86 | -54.21 | 72.48 | 0.3296 |

#### 3.3 SENSITIVITY ANALYSES OF VACCINE EFFICACY

**Table S11. Vaccine efficacy against first *Plasmodium falciparum* infection (Sensitivity Analysis).** VE was estimated as one minus the hazard ratio from a semiparametric proportional hazards model for interval-censored data. Comparisons were made between each vaccine arm and the placebo arm. Profile likelihood 95% confidence intervals and Wald p-values are presented. The protocol-defined primary analysis used a parametric approach; this measure entailed less assumptions on the baseline risk, and is more robust and reliable because the baseline risk was specified according to the placebo.

| Comparison | VE (%) | 95% CI (Lower) | 95% CI (Upper) | P-value (Wald Test) |
|---|---|---|---|---|
| $9x10^5$ PfSPZ Vaccine vs. Normal Saline | 74.12 | 21.30 | 91.49 | 0.0172 |
| $1.8x10^6$ PfSPZ Vaccine vs. Normal Saline | 40.57 | -44.2 | 75.51 | 0.2499 |

**Table S12. Vaccine efficacy based on the proportion of participants with first *Plasmodium falciparum* infection (Sensitivity Analysis).** Vaccine efficacy (VE) was evaluated at 300 days as it is close to the end of study follow-up but precedes the point at which most participants are censored, thus allowing for reliable estimates from the Kaplan-Meier method. The proportion of infection in each treatment arm was estimated as one minus the Kaplan–Meier estimate, assuming infections occurred at the time they were detected. VE was calculated as one minus the relative risk of infection comparing each vaccine arm with the normal saline arm. Exact 95% confidence intervals and p-values were obtained using the melding method implemented in the R package bpcp.

| Comparison | VE (%) | 95% CI (Lower) | 95% CI (Upper) | P-value |
|---|---|---|---|---|
| $9x10^5$ PfSPZ Vaccine vs. Normal Saline | 71.74 | -6.99 | 94.85 | 0.0667 |
| $1.8x10^6$ PfSPZ Vaccine vs. Normal Saline | 48.37 | -51.41 | 83.71 | 0.2763 |

**Table S13. Vaccine efficacy against first Plasmodium falciparum infection stratified on gravidity (Sensitivity Analysis).** Separate baseline hazard functions were specified for each Gravidity stratum while estimating a common treatment effect across strata. Each vaccine group was compared separately with the Normal Saline group. Profile likelihood 95% confidence intervals and Wald p-values are presented.

| Comparison | VE (%) | 95% CI (Lower) | 95% CI (Upper) | P-value (Wald Test) |
|---|---|---|---|---|
| $9x10^5$ PfSPZ Vaccine vs. Normal Saline | 75.13 | 23.24 | 91.94 | 0.0155 |
| $1.8x10^6$ PfSPZ Vaccine vs. Normal Saline | 40.51 | -45.49 | 75.68 | 0.2550 |

**Table S14. Vaccine efficacy against first Plasmodium falciparum infection estimated using an interval-censored proportional hazard model adjusted for birth season (Sensitivity Analysis).** Birth season was included in the model as an adjustment covariate. Separate models were fitted to compare each vaccine arm with the normal saline arm. Profile likelihood 95% confidence intervals and Wald p-values are presented.

| Comparison | VE (%) | 95% CI (Lower) | 95% CI (Upper) | P-value (Wald Test) |
|---|---|---|---|---|
| $9x10^5$ PfSPZ Vaccine vs. Normal Saline | 73.97 | 20.65 | 91.46 | 0.0179 |
| $1.8x10^6$ PfSPZ Vaccine vs. Normal Saline | 39.01 | -48.91 | 75.02 | 0.2776 |

**Table S15. Vaccine efficacy against clinical malaria (Sensitivity Analysis).** VE was estimated as one minus the hazard ratio from a semiparametric proportional hazards model for interval-censored data. Comparisons were made between each vaccine arm and the placebo arm. Profile likelihood 95% confidence intervals and Wald p-values are presented.

| Comparison | VE (%) | 95% CI (Lower) | 95% CI (Upper) | P-value (Wald Test) |
|---|---|---|---|---|
| $9x10^5$ PfSPZ Vaccine vs. Normal Saline | 94.13 | 64.90 | 99.02 | 0.0019 |
| $1.8x10^6$ PfSPZ Vaccine vs. Normal Saline | 35.59 | -53.91 | 73.04 | 0.3223 |

**Table S16. Vaccine efficacy based on the proportion of participants with clinical malaria.** Vaccine efficacy (VE) was evaluated at 300 days as it is close to the end of study follow-up but precedes the point at which most participants are censored, thus allowing for reliable estimates from the Kaplan-Meier method. The proportion of infection in each treatment arm was estimated as one minus the Kaplan–Meier estimate, assuming infections occurred at the time they were detected. VE was calculated as one minus the relative risk of infection comparing each vaccine arm with the normal saline arm. Exact 95% confidence intervals and p-values were obtained using the melding method implemented in the R package bpcp.

| Comparison | VE (%) | 95% CI (Lower) | 95% CI (Upper) | P-value |
|---|---|---|---|---|
| $9x10^5$ PfSPZ Vaccine vs. Normal Saline | 100.00 | 62.50 | 100.00 | 0.0005 |
| $1.8x10^6$ PfSPZ Vaccine vs. Normal Saline | 54.27 | -27.64 | 85.2 | 0.1559 |

**Table S17. Vaccine efficacy against first clinical malaria stratify on gravidity.** Separate baseline hazard functions were specified for each Gravidity stratum while estimating a common treatment effect across strata. Each vaccine group was compared separately with the Normal Saline group. Profile likelihood 95% confidence intervals and Wald p-values are presented.

| Comparison | VE (%) | 95% CI (Lower) | 95% CI (Upper) | P-value (Wald Test) |
|---|---|---|---|---|
| $9x10^5$ PfSPZ Vaccine vs. Normal Saline | 94.46 | 65.63 | 99.11 | 0.0019 |
| $1.8x10^6$ PfSPZ Vaccine vs. Normal Saline | 33.49 | -59.91 | 72.34 | 0.3622 |

**Table S18. Vaccine efficacy against first clinical malaria estimated using an interval-censored proportional hazard model adjusted for birth season.** Birth season was included in the model as an adjustment covariate. Separate models were fitted to compare each vaccine arm with the normal saline arm. Profile likelihood 95% confidence intervals and Wald p-values are presented.

| Comparison | VE (%) | 95% CI (Lower) | 95% CI (Upper) | P-value (Wald Test) |
|---|---|---|---|---|
| $9x10^5$ PfSPZ Vaccine vs. Normal Saline | 94.13 | 64.88 | 99.02 | 0.0019 |
| $1.8x10^6$ PfSPZ Vaccine vs. Normal Saline | 34.50 | -57.14 | 72.70 | 0.3433 |

**Table S19. Protection in the mother during pregnancy was not associated with protection in the infant.**

| Maternal infection status | Vaccine Arm | Infants with Pf infection | Infants without PF infection | % without Pf infection | Infants with CM | Infants without CM | % without CM |
|---|---|---|---|---|---|---|---|
| Mother with Pf infection during pregnancy (n=45) | Normal Saline | 6 | 9 | 60.0% | 6 | 9 | 60.0% |
| | $9x10^5$ PfSPZ | 2 | 14 | 87.5% | 0 | 16 | 100.0% |
| | $1.8x10^6$ PfSPZ | 5 | 9 | 64.3% | 6* | 8 | 57.1% |
| Mother without Pf infection during pregnancy (n=59) | Normal Saline | 4 | 7 | 63.6% | 4 | 7 | 63.6% |
| | $9x10^5$ PfSPZ | 2 | 20 | 90.9% | 1 | 21 | 95.5% |
| | $1.8x10^6$ PfSPZ | 4 | 22 | 84.6% | 4 | 22 | 84.6% |

CM = clinical malaria;

*One clinically ill infant with a positive RDT and negative TBS was classified as CM positive/Pf infection negative using the pre-established case definitions for CM and Pf infection.

Fisher's exact test did not yield any statistically significant differences between groups.

#### 3.4 ANALYSIS OF ANTIBODY RESPONSES

**Table S20. Distribution of OD 1.0 anti-PfCSP IgG Antibodies and Anti-PfMSP1 IgG (ELISA Units) by randomized maternal treatment group and specimen timepoint, with infection statuses combined.**

| Plate Antigen | Treatment group | Timepoint | n | Mean | Median | 25th percentile (Q1) | 75th percentile (Q3) |
|---|---|---|---|---|---|---|---|
| OD 1.0 anti-PfCSP IgG Antibodies | $9x10^5$ PfSPZ | Delivery | 34 | 12,847 | 908* | 539 | 2,866 |
| | | Month 6 | 20 | 355 | 80 | 54 | 144 |
| | | Month 12 | 21 | 130 | 37 | 31 | 102 |
| | $1.8x10^6$ PfSPZ | Delivery | 37 | 2,722 | 1,189* | 496 | 4,384 |
| | | Month 6 | 26 | 294 | 43 | 23 | 127 |
| | | Month 12 | 25 | 119 | 72 | 26 | 126 |
| | Normal Saline | Delivery | 25 | 445 | 344 | 159 | 388 |
| | | Month 6 | 22 | 260 | 45 | 21 | 177 |
| | | Month 12 | 15 | 119 | 92 | 35 | 120 |
| Anti-PfMSP1 IgG (ELISA Units) | $9x10^5$ PfSPZ | Delivery | 34 | 1,245 | 628 | 302 | 1,471 |
| | | Month 6 | 20 | 252 | 10 | 4 | 23 |
| | | Month 12 | 21 | 183 | 4 | 3 | 6 |
| | $1.8x10^6$ PfSPZ | Delivery | 37 | 1,419 | 481 | 193 | 1,051 |
| | | Month 6 | 26 | 182 | 6 | 3 | 11 |
| | | Month 12 | 25 | 685 | 5 | 3 | 366 |
| | Normal Saline | Delivery | 25 | 1,031 | 453 | 241 | 1,043 |
| | | Month 6 | 22 | 290 | 7 | 3 | 12 |
| | | Month 12 | 15 | 1,545 | 19 | 3 | 927 |

*indicates significant difference compared to normal saline at the same timepoint ($p<0.0001$) using Nonparametric, two-sided Mann–Whitney U tests. P-values were not corrected for multiple comparisons.

#### 4. SUPPLEMENTARY FIGURES

**Figure S1. PfCSP-specific IgG antibody responses measured by ELISA in vaccinated mothers at delivery and in their infants at 6 and 12 months of age.** Data are presented as the median and interquartile range of the reciprocal serum dilution at which the OD was 1.0 and are shown separately for participants receiving 9.0 × $10^5$ PfSPZ Vaccine, 1.8 × $10^6$ PfSPZ Vaccine, and normal saline controls. Within each panel, responses are stratified by infant infection status during the corresponding follow-up period. Each symbol represents an individual participant; horizontal lines indicate medians, and error bars represent interquartile ranges. OD 1.0 values are displayed on a $\log_{10}$ scale. Of the 104 infants followed (38 in 9x105; 40 in 1.8x106; 26 in the control group) 34/38, 37/40 and 25/26 of the mothers had sera available at the time of delivery, 20/38, 26/40, and 22/26 of the infants had sera available at 6 months and 21/38, 25/40, and 15/26 of the infants had sera available at 12 months. Differences between groups were assessed using the Mann-Whitney U test. The p-values were not corrected for multiple comparisons.

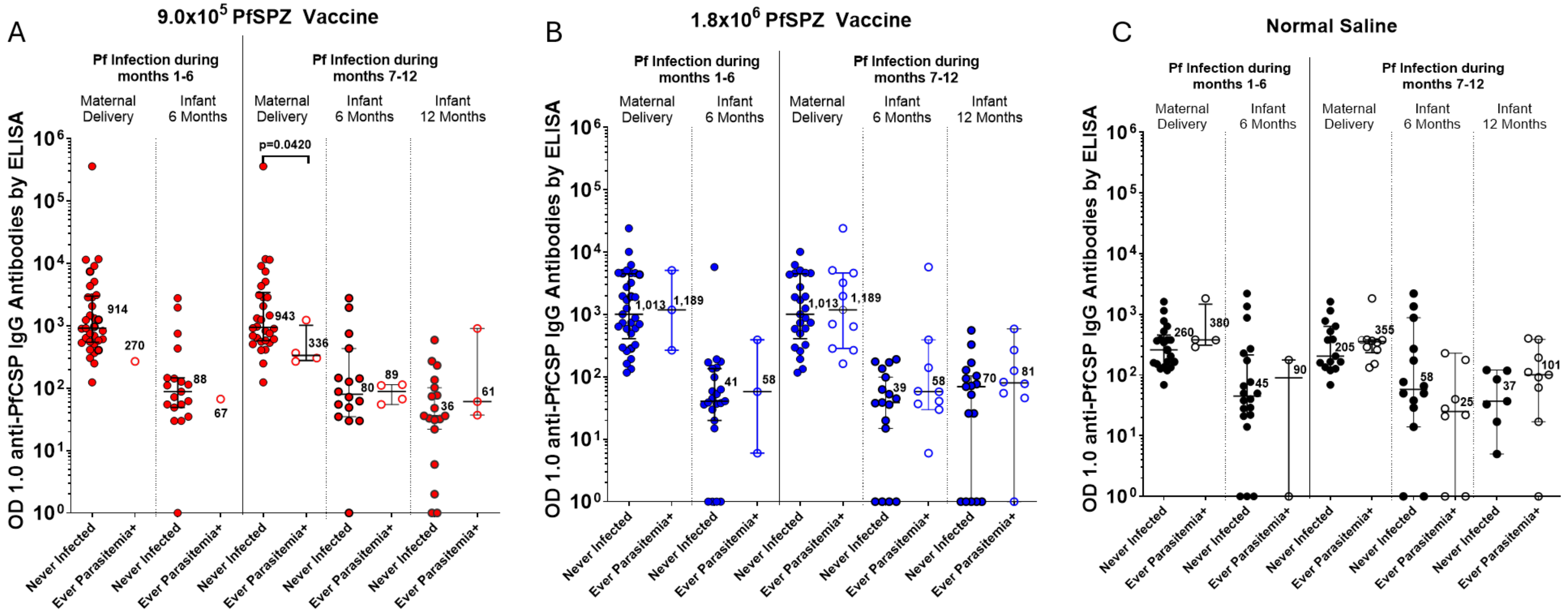

**Figure S2. PfMSP1-specific IgG antibody responses measured by ELISA in vaccinated mothers at delivery and in their infants at 6 and 12 months of age.** Data are presented as the median and interquartile range of anti-PfMSP1 ELISA units (EU) and are shown separately for participants receiving 9.0 × $10^5$ PfSPZ Vaccine, 1.8 × $10^6$ PfSPZ Vaccine, and normal saline controls. Within each panel, responses are stratified by infant infection status during the corresponding follow-up period. Each symbol represents an individual participant; horizontal lines indicate medians, and error bars represent interquartile ranges. ELISA units are displayed on a $\log_{10}$ scale. The horizontal reference lines indicate the respective PfMSP1 ELISA limit-of-detection (LoD) seropositive thresholds for maternal and infant samples. Of the 104 infants followed (38 in 9x$10^5$; 40 in 1.8x$10^6$; 26 in the control group) 34/38, 37/40 and 25/26 of the mothers had sera available at the time of delivery, 20/38, 26/40, and 22/26 of the infants had sera available at 6 months and 21/38, 25/40, and 15/26 of the infants had sera available at 12 months. Differences between groups were assessed using the Mann-Whitney U test. The p-values were not corrected for multiple comparisons.

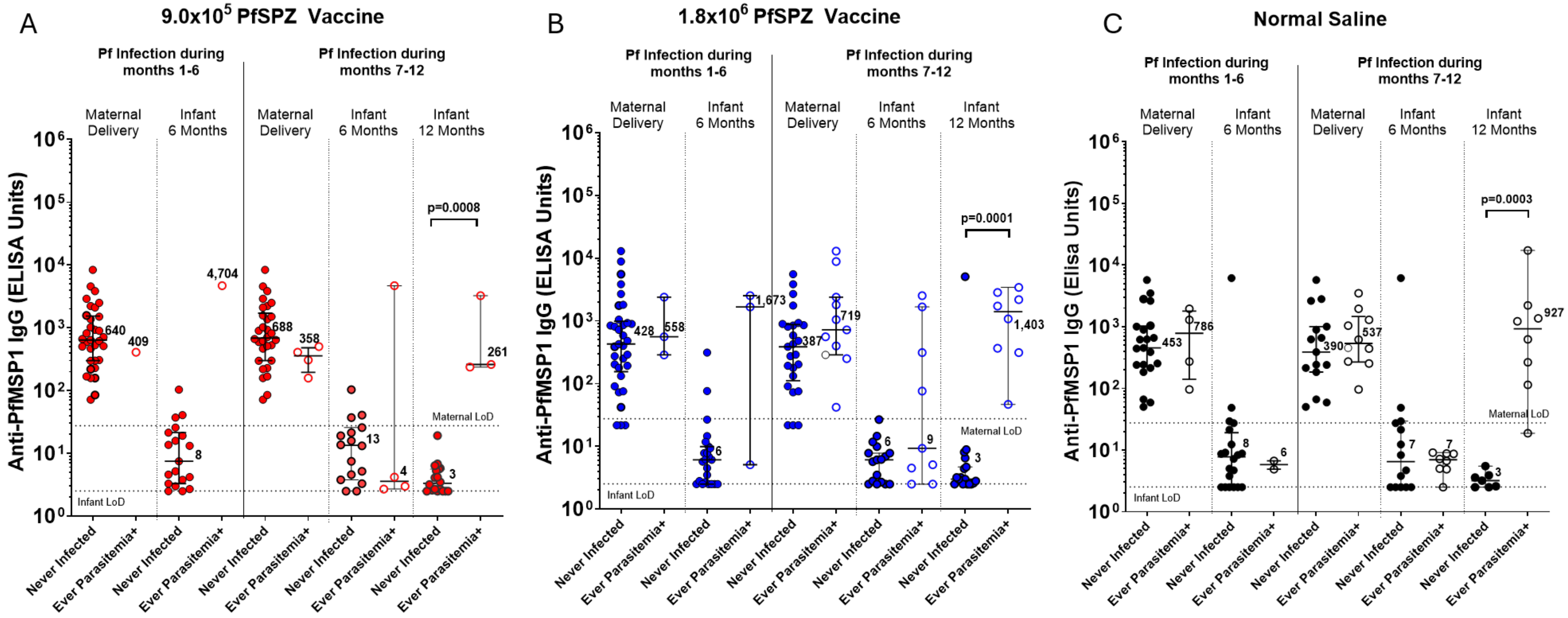

**Figure S3. γδ Vδ2 T cell levels in maternal delivery or infant samples**

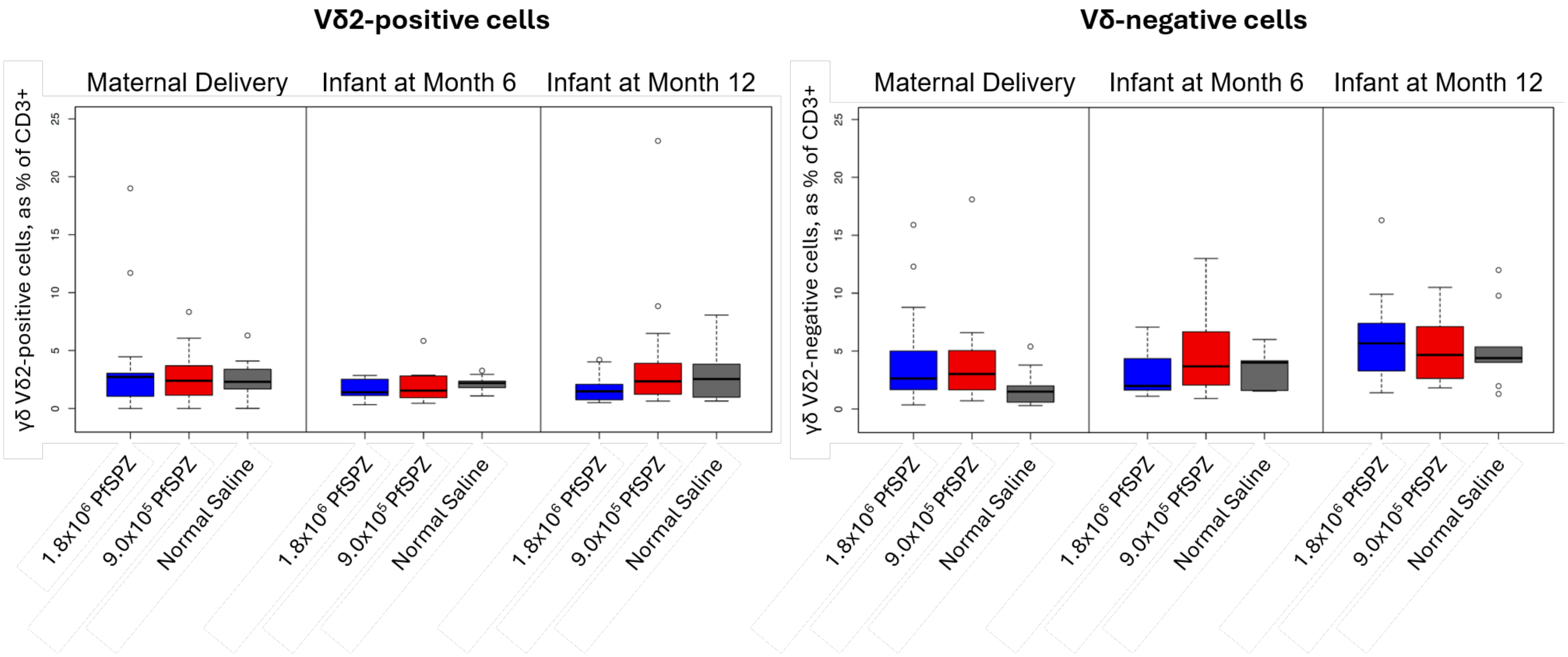

**Figure S4. γδ Vδ2 T cell fold change in infant samples from maternal delivery sample baseline**

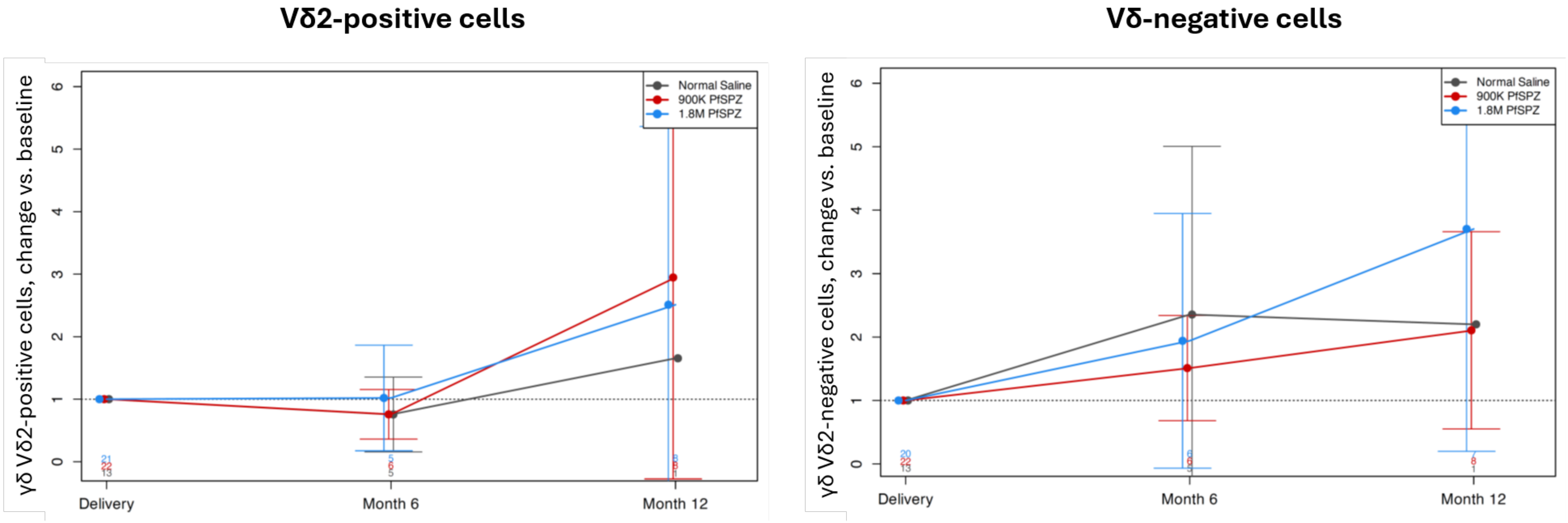
